# Training-free Design of Deep Networks as Ensembles of Clinical Experts

**DOI:** 10.1101/2024.03.17.24304438

**Authors:** Tinghui Wu, Jieke Wu, Zijun Zhang, Wuyang Chen

## Abstract

Artificial intelligence (AI) techniques such as deep learning hold tremendous potential for improving clinical practice. However, clinical data complexity and the need for extensive specialized knowledge represent major challenges in the current, human-driven model design. Moreover, as human interpretation of a clinical problem is inherently encoded in the model, the conventional single model paradigm is subjective and cannot fully capture the prediction uncertainty. Here, we present a fast and accurate framework for automated clinical deep learning, TEACUP (training-free assembly as clinical uncertainty predictor). The core of TEACUP is a newly developed metric that faithfully characterizes the quality of deep networks without incurring any cost for training of these networks. When compared to conventional, training-based approaches, TEACUP reduces computation costs by more than 50% while achieving improved performance across distinct clinical tasks. This efficiency allows TEACUP to create ensembles of expert AI models, contributing to recommendations in clinical practice by mimicking the approach of using multiple human experts when interpreting medical data. By combining multiple perspectives, TEACUP provides more robust predictions and uncertainty quantification, paving the way for more reliable clinical AI.

## 1 Introduction

Deep neural networks (DNNs) have provided state-of-the-art tools for many machine learning tasks in clinical and biomedical applications, including disease classification [1, 2], medical imaging segmentation [3], medical device signal processing [4, 5], and genetic variant pathogenicity prediction [6, 7]. These clinical data modalities vastly differ from each other, making the construction of DNN models handling specific clinical datasets expertise-dependent. In routine clinical care, computed tomography (CT) scans, capturing detailed internal body structures in 3D, are typically modeled using 3D-convolutions, whereas electrocardiogram (ECG) recordings, which represent temporal dynamics of heart electrical activity in 1D, necessitate 1D-convolution models. Similarly, breast ultrasound images, commonly used to detect abnormalities such as benign or malignant tumors, are processed as 2D images and can be modeled effectively using 2D-convolutions to capture spatial features in the image that aid in diagnosis. Furthermore, the design of high-quality DNN models, even for the same clinical data modality, can vary significantly. For instance, different DNNs were designed to analyze and interpret ECG data to aid in the diagnosis of abnormal heart rhythms [4] versus asymptomatic left ventricular dysfunction [8]. Overall, designing clinical DNN models currently requires a deep knowledge of medicine, an expertise in deep learning techniques, and time to manually perform many trial and error analyses. Therefore, developing DNN design strategies that don’t depend on empirics-driven model design would eliminate these limitations and offer more scalable and data-driven solutions for clinical artificial intelligence (AI).

Automated machine learning (AutoML) algorithms, unlike conventional machine learning [9–11], can overcome the dependence on empirics-driven expertise by automating the design of high-quality models. Conventionally, AutoML methods aim to provide a one-size-fits-all solution that can be applied to various datasets and prediction tasks, and are designed to work with both conventional machine learning and deep learning models. TPOT, a popular tool in this category, is a tree-based pipeline optimization tool that leverages feature preprocessors and model architectures to achieve optimal performance [12, 13]. AutoML methods tailored to DNNs have recently been used to design DNNs in specific domains, tasks, or types of data. For example, the Automated Modelling for Biological Evidence-based Research (AMBER) framework uses AutoML algorithms and Neural Architecture Search (NAS) to design DNN models that classify genomic sequences and identify disease-relevant genetic variants [14, 15].

However, all these AutoML strategies require model training to estimate the quality of the designed networks [16], which can compromise the efficiency and accuracy of the model search [17, 18]. DNN training is too time-consuming and computationally costly, routinely requiring hundreds [19] to dozens of thousands [20] of GPU hours; thus, the use of AutoML methods for clinical DNNs in the broader clinical and biomedical community has been prohibitive.

To overcome these obstacles, we present TEACUP (Training-free Assembly as Clinical Uncertainty Predictors), an automated deep learning framework for the rapid and accurate design of DNNs that tackle clinical tasks. TEACUP is modular and effectively separates the automated DNN design into evaluation and search phases (Fig. 1). During the evaluation phase, TEACUP leverages a novel composite training-free metric that characterizes a model’s performance without any model training. TEACUP then optimizes the composite training-free metric to search for high-quality, task-specific DNN architectures, compatible with a wide range of searching algorithms. Importantly, the TEACUP architecture ensemble quantifies prediction uncertainty (Fig. 1B), which is not feasible in the human-expert design or training-based AutoML design of neural networks, due to prohibitive computation costs. In clinical applications, uncertainty quantification can help clinicians make more informed and accurate decisions. For example, uncertain AI predictions can serve as a trigger for additional clinical investigations and more comprehensive, orthogonal clinical tests. On the contrary, highly confident AI predictions may reduce unnecessary testing or interventions. Indeed, multiple computational methods must agree on the same prediction before they can be considered clinically [21]. Thus, TEACUP uniquely allows clinicians to prioritize high-confidence AI predictions and to flag uncertain predictions for further clinical investigation. This key distinction leads to more accurate diagnoses, fewer unnecessary interventions, and improved patient outcomes [22].

**Fig. 1:**
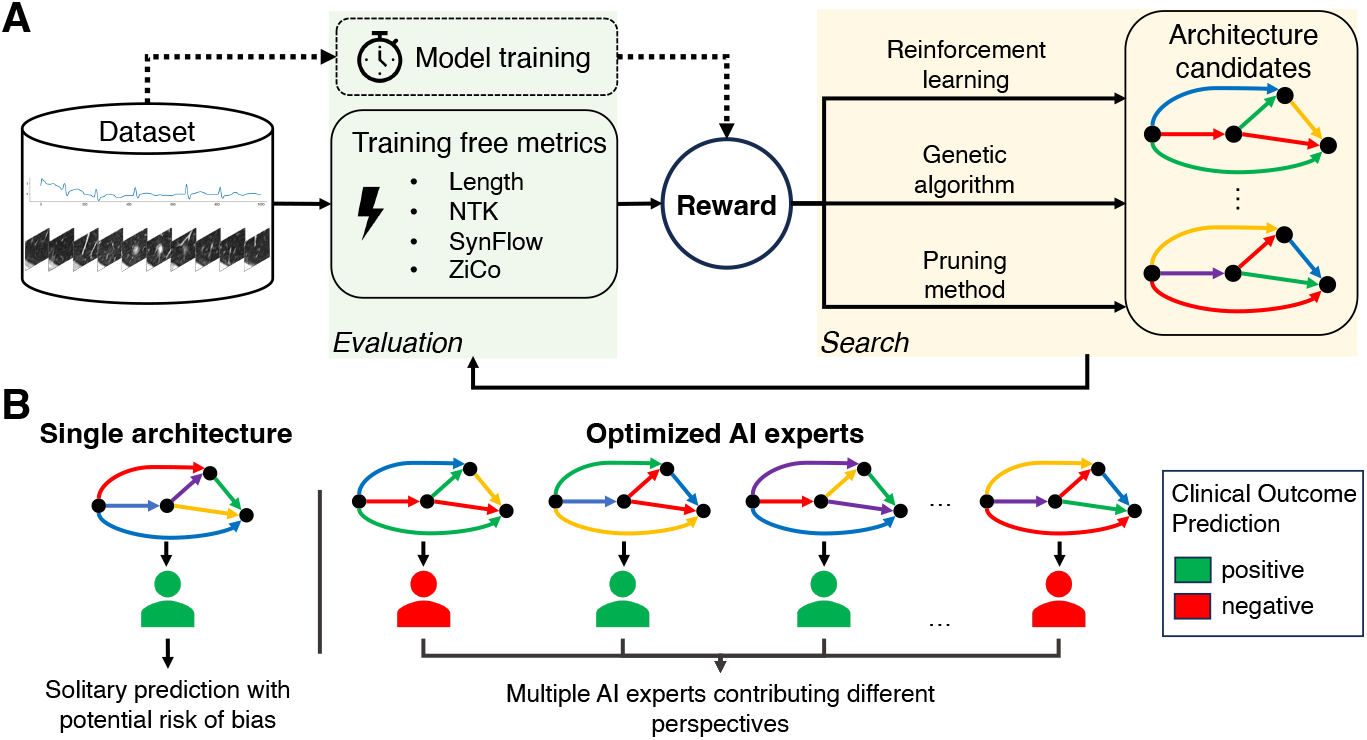
TEACUP enables fast model evaluation on diverse clinical tasks with training-free metrics. **(A)** The workflow of the TEACUP framework. Instead of relying on traditional model training for DNN design, the reward of each sampled architecture is generated from the TEACUP metric, a weighted combination of calculated training-free metrics. Then, search algorithms optimize architectures based on rewards. **(B)** Illustration of the conventional single model architecture prediction and an ensemble of distinct models designed by our TEACUP. Compared to deep ensemble of AI experts, the single model often cannot fully capture the prediction uncertainty, posing potential risk of bias.

## 2 Results

### TEACUP characterizes model performance across clinical tasks without training

The core innovation of TEACUP is to develop and apply training-free methods that can faithfully evaluate a DNN’s quality, without actually training the model. This is facilitated by previous extensive theoretical and empirical studies on natural images (e.g. CIFAR-100). However, clinical tasks harbor distinct challenges compared to natural images. We systematically introduced four training-free metrics: (1) Length distortion (Length), which measures the extent to which the network distorts an input curve [23–25]; (2) condition number of Neural Tangent Kernel (NTK), which indicates the trainability of a neural network [26–28]; (3) Synaptic flow (SynFlow), which is based on DNN parameter pruning and measures the sensitivity to changes in a specific parameter [29, 30]; and (4) Zero-shot Inverse Coefficient (ZiCo), which jointly considers the mean and standard deviation of a DNN’s sample-wise gradients for high training convergence speed and generalization capacity [31] (Methods 7.2). Therefore, these four training-free metrics characterize the properties of a DNN in a complementary fashion and, taken together, provide a promising way of evaluating a DNN.

To rigorously evaluate the quality of these training-free metrics, we designed a comprehensive benchmark from clinical task and model architecture perspectives. First, from clinical task perspective, we performed three distinct clinical tasks with disparate application and data dimensions. Specifically, we used TEACUP to generate models from 1D ECG recordings for multi-classification of atrial fibrillation (ECG2017) [15], 2D ultrasound images for detection of breast abnormalities (BreastMNIST) [32], and 3D CT scan images to classify cancerous lung nodules (NoduleMNIST3D) [33] (see Methods 7.1 for details). Second, from model architecture perspective, we diversely sampled *n* = 150 models from the NAS-Bench-201 model space, one of the most popular and powerful deep residual convolutional model spaces widely adopted in deep learning [34] (see Supplementary Figure 1 for an illustration). NAS-Bench-201 model space provides a predefined set of neural network architectures, and we evaluated the corresponding performance metrics on each specific benchmark dataset. The sampled models correspond to approximately 1% of all possible models (Methods 7.3). We then compared the models ranked by training-free metrics to the performances of models that were fully trained to convergence. Specifically, we measured the Spearman correlation coefficient (*ρ*) of training-free metrics with the testing performance on datasets for ECG2017 (F1 score), BreastMNIST (AUC score), and NoduleMNIST3D (AUC score) (Fig. 2; Supplementary Table 2).

**Fig. 2:**
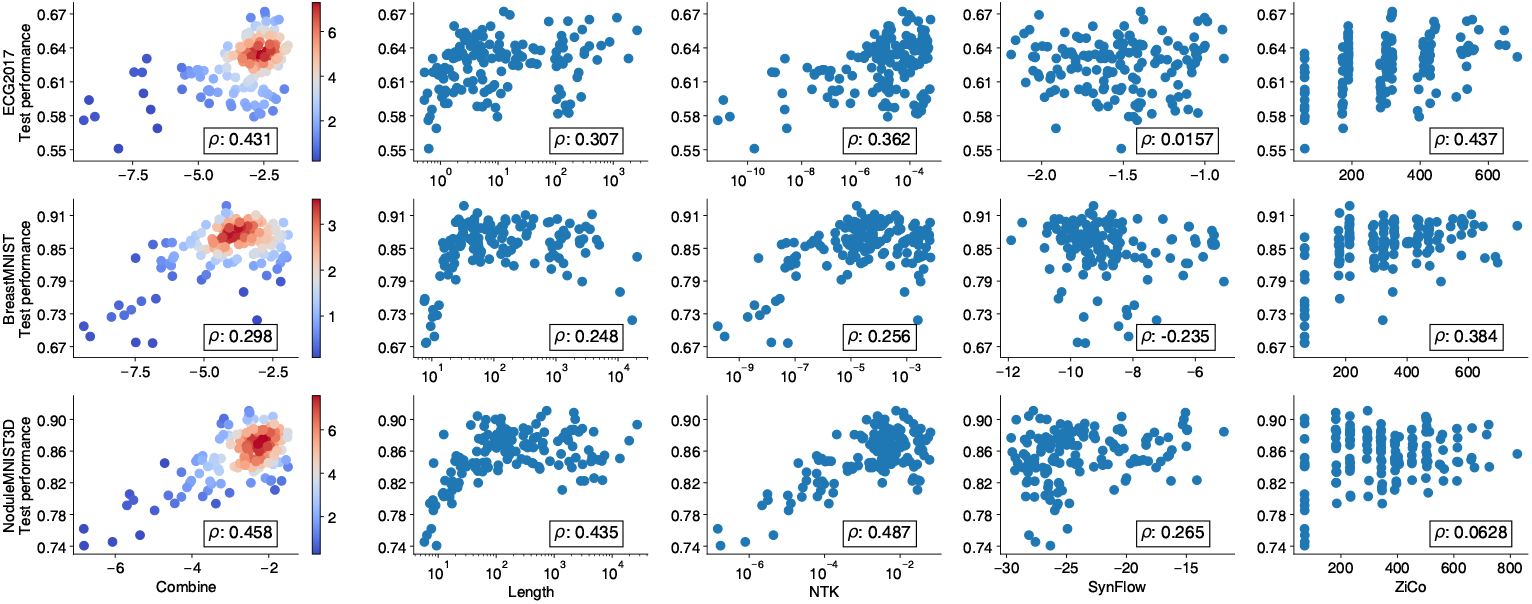
Spearman correlation between the test reward and training-free metrics. A composite TEACUP metric (left) is derived by combining all four metrics: Length, NTK, SynFlow, ZiCo.

Importantly, no single existing training-free metric can consistently outperform other metrics across ECG2017, BreastMNIST, and NoduleMNIST3D datasets. Each of the four training-free metrics provided different levels of information when compared to their fully-trained model. While ZiCo achieved superior performance in ECG2017 (*ρ* = 0.437), it failed to capture the trained performance in NoduleMNIST3D (*ρ* = 0.063). By contrast, NTK performed substantially better in NoduleMNIST3D (*ρ* = 0.487) than in ECG2017 (*ρ* = 0.362).

Motivated by the different behaviors and complementary predictions of these four training-free metrics, we sought to develop a new composite training-free metric that can consistently characterize a DNN’s quality across different clinical tasks. We refer to this new composite metric as the TEACUP metric that combines all four existing training-free metrics. We performed a multivariate linear regression analysis to find the most predictive linear combination of training-free metrics (Methods 7.4) on ECG2017 and NoduleMNIST3D. Even though test rewards in ECG2017 and NoduleMNIST3D datasets differ by a scale factor, our bootstrapped regression analysis revealed a surprisingly simple combination: TEACUP metric= *−*0.5× log(Length) + log(NTK) + log(SynFlow) + log(ZiCo) (Supplementary Figure 2). We directly adopt this combination on the BreastMNIST dataset. As shown in Fig. 2, the composite TEACUP metric that combines all training-free metrics, Length, NTK, SynFlow, and ZiCo, consistently achieved an overall high Spearman correlation with the performance of trained models (*ρ* = 0.471 for ECG2017, *ρ* = 0.298 for BreastMNIST, *ρ* = 0.442 for NoduleMNIST3D). We further demonstrated that all four training-free metrics, despite their various level of correlations to model performance in clinical tasks, were all informative to the final performance of the TEACUP metric (Supplementary Table 3).

### TEACUP significantly reduces computing time and costs

We quantified the computing costs saved by employing the TEACUP metric versus training DNN models. To set up the comparison, we tested a commonly used, naive baseline approach that assumes partially-trained models (i.e., only trained for the first few epochs) can predict DNN architecture generalization, therefore providing a fair comparison to our TEACUP metric performance. To determine how many training samples were consumed to achieve the same correlation as the TEACUP metric in each clinical dataset, we measured the correlation between the validation performance (F1 for ECG2017, AUC for BreastMNIST and NoduleMNIST3D) at each trained epoch and the test performance across *n* = 150 models. Notably, the correlation coefficient, when computed against the validation performance from each trained epoch across all *n* = 150 architectures, will be close to, but not reach 1. This is expected because even though there is a strong relationship between the validation and test rewards, they are not perfectly aligned.

As shown in Fig. 3, on a single A100 GPU each model will take almost 10 minutes to train on ECG and lung nodule scans to reach the same correlation to the model’s test performance as the TEACUP metric, corresponding to training on roughly 300,000 samples in ECG2017 and 35,000 samples in NoduleMNIST3D. In contrast, computing the training-free TEACUP metric takes less than 10, 10, and 40 seconds to achieve a high correlation on ECG2017, BreastMNIST, and NoduleMNIST3D, respectively. This represents a substantial reduction of over 50% in computing cost every time a DNN model is evaluated. Equivalently, using TEACUP framework on the scale of a previous milestone study that evaluated 12,800 architectures [20], would be predicted to save over $7,300 in cloud computing costs and 1,792 hours per AutoML experiment ^1^.

**Fig. 3:**
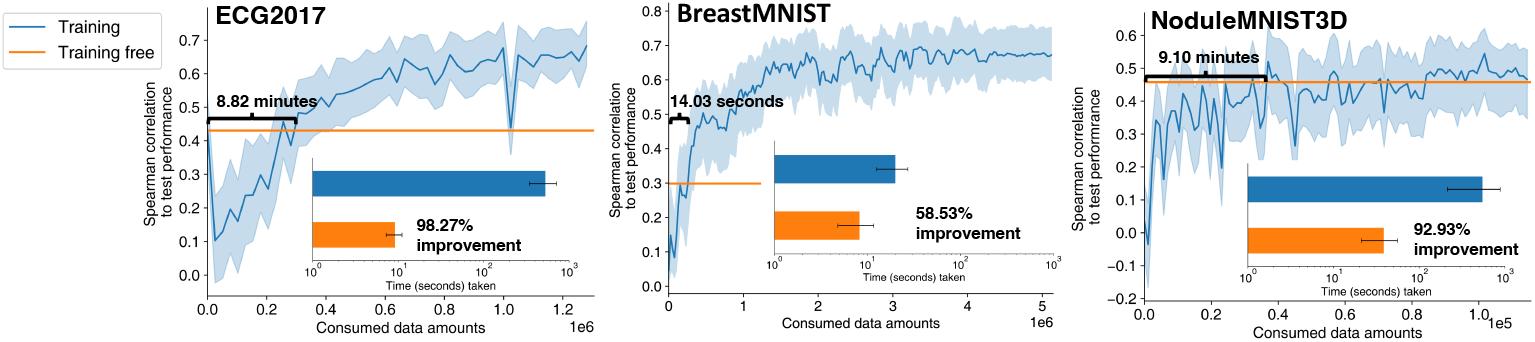
Comparison of computation time costs between partial training and the TEACUP metric when they achieve equal Spearman correlations with the final trained DNN performance. For “Training”, the Spearman correlation across the 150 benchmark architectures (y-axis) is calculated between the testing and the validation accuracy after trained for a certain amount of data (x-axis). Orange horizontal lines correspond to values of *ρ* in the left column of Fig. 2.

### TEACUP improves the accuracy of automated model design

We systematically evaluated three distinct searching algorithms to optimize neural architectures using the TEACUP metric as optimization objectives. In particular, we implemented a reinforcement learning-based controller network [14, 20] and a probabilistic model building genetic algorithm [35] to maximize the TEACUP metric as a reward, and a pruning-based algorithm that iteratively removes the edge with the minimum contribution to the TEACUP metric from a super-net [28]. Given that these three algorithms have distinct underlying assumptions for DNN architectures and cover a wide range of neural architecture search methods, their performance would be indicative of the general utility of the TEACUP metric in automated DNN design. TEACUP removed heavy computation costs in conventional AutoML methods and allowed us to efficiently perform 10 independent runs for each search algorithm on each clinical task, which enabled us to account for the stochasticity of the search algorithms. This would not have been feasible using conventional AutoML methods for DNN, given the cost and time required to finish these tasks. In contrast, all three search algorithms were efficient in optimizing the TEACUP metric and converged within 6 GPU hours across all runs with TEACUP (Methods 7.5; Supplementary Figure 3).

The improved TEACUP metrics successfully translated to high-performance DNN models. To examine the quality of DNN models built using TEACUP, we first compared TEACUP-optimized models to randomly sampled models from the NAS-Bench-201 model space, then expanded the comparison to state-of-the-art AutoML methods. TEACUP-optimized models exhibited significantly higher testing performance in all three datasets, with the sole exception of pruning-based search method in ECG2017 which failed to improve performance (Fig. 4). Nonetheless, in NoduleMNIST3D dataset, pruning-based algorithm searched for models (test AUC=0.88 ± 0.02; Table 1) significantly better than the randomly sampled benchmark (test AUC=0.85 ± 0.03). This demonstrates that different search algorithms have varying performances across clinical datasets. Indeed, we found that the reinforcement learning-based controller network achieved superior searched model performance in the ECG2017 dataset, while genetic algorithm worked the best in the NoduleMNIST3D dataset, and all three search algorithms largely outperformed random search on BreastMNIST. Furthermore, these TEACUP-optimized models are significantly more accurate than previously published human-designed baselines (Table 1). In ECG2017 [15], TEACUP models achieved an average testing F1 of 0.66 ± 0.01, significantly higher than XGBoost (0.44 ± 0.02) [36] and Wide ResNet (0.57 ± 0.01) [37]. Similarly in NoduleMNIST3D [33], TEACUP models performed better (testing AUC of 0.89 ± 0.01) than ResNet-18 (0.88 ± 0.02) and a substantially more parameterized ResNet-50 (0.87 ± 0.02) [38]. Even compared with training-based AutoML methods [14, 39–42], TEACUP remained highly competitive and matched the performance among state-of-the-art AutoML methods. Additionally, we ensembled the TEACUP models into the TEACUP ensemble (Fig. 4), which achieved the highest test performance among existing AutoML methods (test F1=0.68 on ECG2017, test AUC=0.94 on BreastMNIST, test AUC=0.92 on NoduleMNIST3D).

**Table 1:** Performance comparison between our approach and previous works. “RL”: reinforcement learning. “Genetic”: genetic algorithm.

| Dataset | ECG2017 |  | BreastMNIST |  | NoduleMNIST3D |  |
| --- | --- | --- | --- | --- | --- | --- |
| | Method | Result (F1)<br>Mean ± Std (Ensemble) | Method | Result (AUC)<br>Mean ± Std (Ensemble) | Method | Result (AUC)<br>Mean ± Std (Ensemble) |
| Training-free<br>searching method | RL | $0.66 \pm 0.01$ (**0.68**) | RL | $0.89 \pm 0.01$ (**0.93**) | RL | $0.87 \pm 0.01$ (0.90) |
| | Genetic | $0.65 \pm 0.01$ (**0.68**) | Genetic | $0.90 \pm 0.01$ (**0.93**) | Genetic | $0.89 \pm 0.02$ (**0.92**) |
| | Pruning | $0.62 \pm 0.04$ (0.66) | Pruning | $0.86 \pm 0.05$ (0.92) | Pruning | $0.88 \pm 0.02$ (0.91) |
| Fixed architecture<br>training | Wide ResNet (WRN) | $0.57 \pm 0.01$ | ResNet-18 | $0.88 \pm 0.02$ | ResNet-18 | $0.88 \pm 0.02$ |
| | XGBoost | $0.44 \pm 0.02$ | ResNet-50 | $0.87 \pm 0.02$ | ResNet-50 | $0.87 \pm 0.02$ |
| Training-based<br>searching method | DenseNAS random | $0.58 \pm 0.01$ | Auto-sklearn | $0.84 \pm 0.02$ | Auto-sklearn | $0.91 \pm 0.01$ |
| | DARTS GAEA | $0.66 \pm 0.01$ | AutoKeras | $0.83 \pm 0.03$ | AutoKeras | $0.87 \pm 0.03$ |
| | AMBER | $0.67 \pm 0.02$ | | | | |

**Fig. 4:**
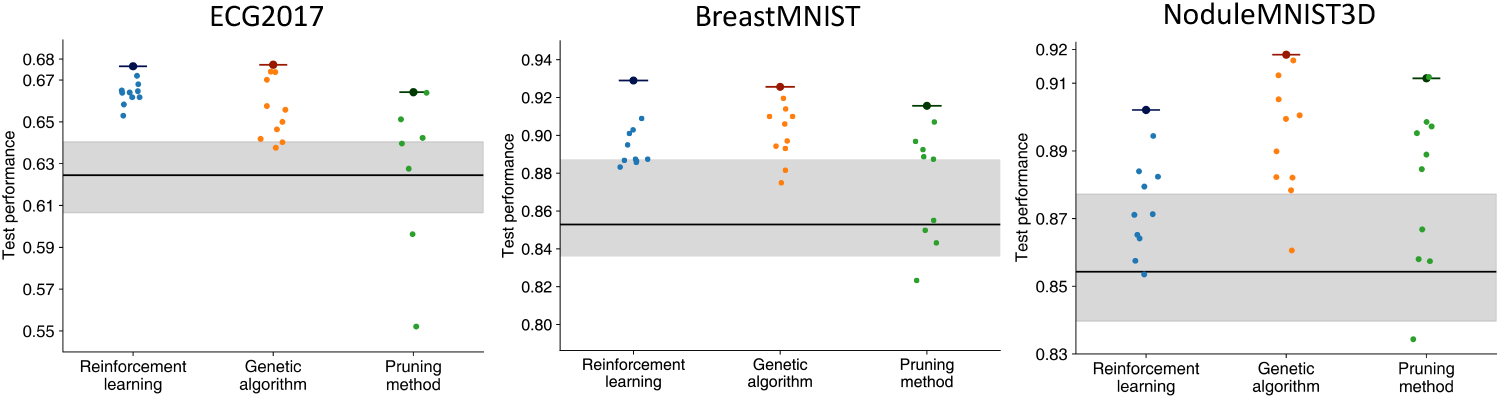
Performance of ensemble prediction (larger dot with a dash) and optimized architectures (dot) compared with benchmark results (gray area). The horizontal line is the median, and the gray area is the upper to lower quartile region, for the performance of 150 benchmark architectures, respectively.

The automated TEACUP-optimized DNNs not only achieved strong accuracy but also discovered task-specific patterns of DNN structures that are aligned with characteristics of different tasks and clinical data modalities (Fig. 5). In the case of ECG2017, more edges are allocated with pooling layers and convolutions with the larger kernel size. In contrast, skip connections and the small convolution kernel are preferred in NoduleMNIST3D. These different groups of architectures are highly aligned with the characteristics of clinical datasets. In ECG2017, the long sequence of the heartbeat signal (1000 in length) requires a larger receptive field, which can be achieved by pooling and large kernels. However, 3D images of lung nodule scans have a much smaller spatial resolution (28 × 28 × 28), and using small convolutions will achieve sufficient receptive fields.

**Fig. 5:**
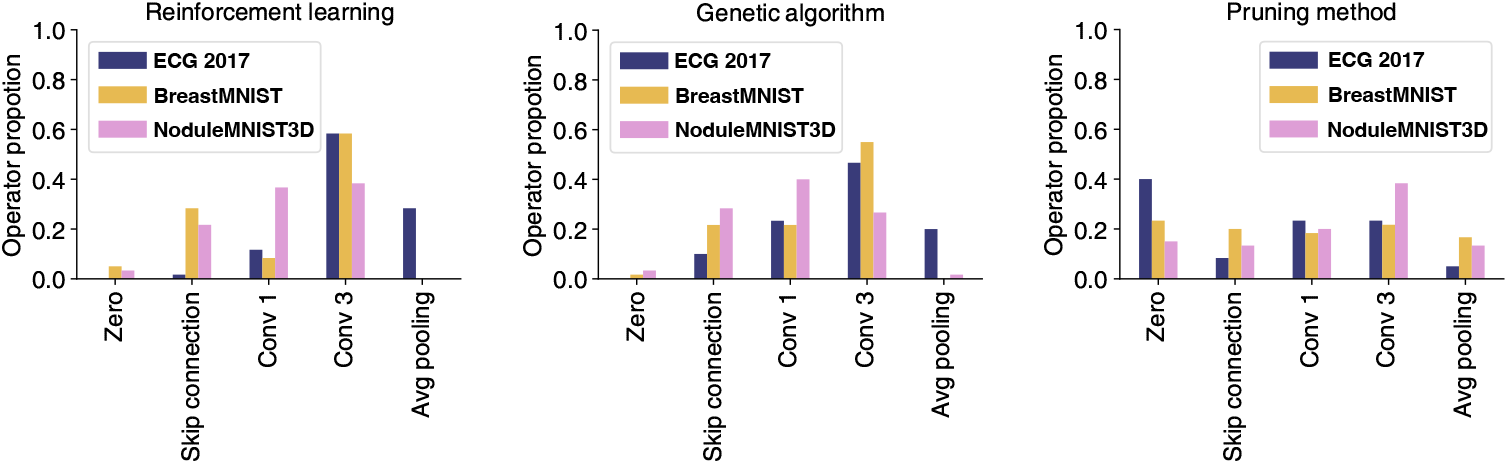
Distributions of optimized architectures by reinforcement-learning, genetic-algorithm, and pruning-method on three datasets.

### TEACUP enables robust ensemble learning and uncertainty quantification

The multiple accurate, distinct architectures efficiently optimized by TEACUP present an unprecedented opportunity for uncertainty quantification, which is impossible to achieve using conventional single-model paradigms given the prohibitive DNN training cost. In the context of DNNs, different types of neural networks can make incorrect clinical predictions for the same patient persistent to an architecture, even when starting from different weight initializations, suggesting that each neural network is sensitive to different aspects of the data [43].

We hypothesized that DNNs of different architectures optimized by TEACUP capture different perspectives of clinical data patterns and thus would allow us to prioritize high-confidence, high-accuracy predictions over uncertain, less-accurate predictions. To rank predictions based on their confidence, we computed their variance across different TEACUP-optimized models as a measure of uncertainty quantification (Method 7.7). Intuitively, neural network models with distinct architectures (i.e. different “experts”) that make the same prediction from clinical samples are more confident and thus likely to be accurate. For comparison, we followed the same uncertainty computation across different random initializations of a single model architecture, which already goes beyond the conventional single model regime by considering multiple random weight initializations. In the test NoduleMNIST3D example illustrated in Fig. 6A, even though the predictions from both schemes are incorrect (i.e. false negative), the architecture-wise uncertainty calculated by the TEACUP ensemble is high, flagging this example for additional tests; whereas the initialization uncertainty calculated by the single-architecture ensemble is low, indicating over-confidence in its incorrect prediction.

**Fig. 6:**
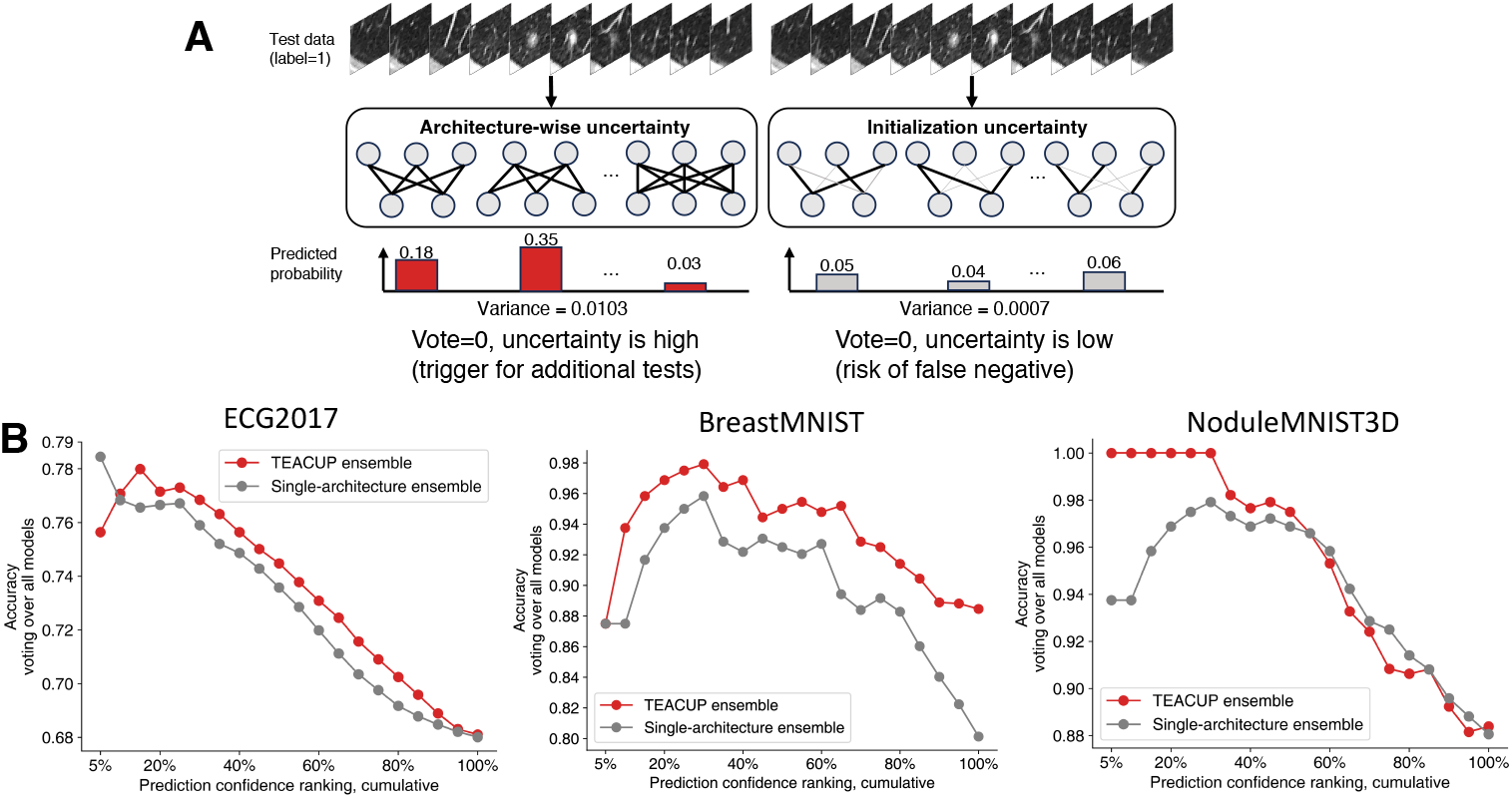
**(A)** Comparison of a false negative test case over two uncertainty quantification methods: different architectures vs. single architecture trained from different weight initialization. Despite both methods yielding incorrect predictions, the larger architecture-wise uncertainty suggests the need for additional tests. **(B)** Across testing samples of different confidence, the ensemble of TEACUP-optimized models (search method: RL for ECG2017, pruning for BreastMNIST, Genetic for NoduleMNIST3D) outperforms the ensemble of a single architecture trained from random initialization.

We further chose the architecture that occurred most frequently across all TEACUP search runs as the single model architecture baseline for ECG2017, BreastMNIST, and NoduleMNIST3D (Method 7.7). This represents a strong and fair baseline, because the DNN architecture that is consistently searched as optimal by all three search algorithms is more likely to fit the corresponding clinical data. Consequently, this allowed us to interrogate the influence of assembling distinct architectures versus only weight initialization variability in an optimal but fixed architecture.

Deep ensembles of TEACUP-optimized distinct architectures showed superior performance in prioritizing high-confidence clinical predictions, even when compared against the strong baseline of the most consistently-searched single architecture ensemble. Although both methods were highly competitive when evaluating all test samples (rightmost dot, 100% test data; Fig. 6B), the TEACUP ensemble achieved a superior accuracy compared to the single-architecture ensemble when we move to the most highconfidence predictions, (leftmost dots, Fig. 6B). For example, the TEACUP ensemble achieved perfect accuracy (100%) in 30% of top-ranked confident predictions in NoduleMNIST3D; whereas the single-model ensemble achieved an accuracy of only 94%, highlighting a 6% accuracy gap. Thus, the TEACUP ensemble more effectively ranks and prioritizes high-confidence, high-accuracy predictions.

## 3 Discussion

In this study, we presented TEACUP, an automated workflow for designing deep learning models tailored to clinical tasks. By using training-free metrics, we can estimate a neural network’s performance without expending large computation resources on model training, reducing computation cost by more than 50% per model evaluation. The composite metric we developed is compatible with a wide range of search algorithms. We demonstrated improved training-free metrics using three search algorithms. With the evaluation of three distinct datasets varying in dimension and application, TEACUP yielded promising results, showing its potential to adapt to a variety of clinical tasks. Foundation models have been a rising trend and adapting to the medical domain as well, with widespread adoption in various downstream tasks, due to their exceptional performance and versatility [44, 45]. However, despite their efficacy, it is also known that building foundation models requires huge amounts of data and high computation resources for training. Furthermore, a significant challenge lies in the interpretability of foundation models, particularly crucial in clinical applications where understanding the decision-making process is important. In contrast, TEACUP offers an alternative solution that not only accelerates the model development process but also facilitates the exploration of multiple architectural choices, mitigating the risk of bias associated with relying solely on a single model. By analyzing the distribution among different model predictions, our approach provides insights into the overall uncertainty of the results. Additionally, as our models leverage well-established deep learning frameworks, a valuable future direction is to utilize explainable AI tools like Grad-CAM to understand the model’s decision-making process by elucidating the elements of input data.

There are a couple of areas for improvement in our approach. The first one lies in the selection of the model space. NAS-Bench-201, the model space we utilized, has shown excellent performance on the famous natural image classification dataset, CIFAR-100. However, it remains unknown whether this model space spans the best space to represent all the possibilities of model architecture for medical data analysis. Future research could explore alternative model spaces specifically tailored to medical datasets, considering the unique characteristics and complexities of medical data. Another notable feature is that TEACUP only requires a small portion of the data for robust training-free metrics calculation, which holds considerable promise in medical data analysis where data is scarce. Therefore, future investigations would focus on refining data subset selection methodologies, which has the potential to significantly enhance the efficiency and applicability in clinical practice.

In summary, TEACUP is a fast, accurate, and universal automated deep learning framework that could be easily adapted to different clinical tasks with light computation resources. TEACUP generated multiple, optimal models with distinct architectures that enabled architecture-wise ensemble learning, which in turn allowed us to quantify DNN uncertainty for clinical data for the first time. As such, the neural uncertainty enabled by our TEACUP framework presents a major paradigm shift to current deep learning-based clinical prediction methods, and we expect that it will be of significant clinical utility.

## 4 Data Availability

The benchmarking datasets are publicly available. ECG2017 is precompiled in https://nb360.ml.cmu.edu/. Both BreastMNIST CT scans and Lung nodule scan is part of the MedMNIST dataset in https://medmnist.com/.

## 5 Code Availability

Our code and analysis can be found in GitHub: https://github.com/zhanglab-aim/TEACUP.

## 6 Acknowledgements

We acknowledge all members of the Zhang lab for helpful discussions.

## 7 Methods

### 7.1 Data

To demonstrate the performance of our method, we study three benchmarks for machine learning tasks routinely used in clinical settings, covering 1D, 2D, and 3D medical data:

1. ECG (Electrocardiogram): The dataset [15] is originally collected for the 2017 PhysioNet Challenge [46] for heartbeat categorization, with an average 30-second ECG recording stored at 300 Hz. Processed by 1,000 ms sliding window and 500 ms stride, there are 261,740 training data, 33,281 validation data, and 33,494 testing data in total. All the data is labeled from one of the four classes: normal, disease (atrial fibrillation, AF), other, or noisy rhythm. The macro-averaged F1 score is used to evaluate training performance.
2. BreastMNIST: This dataset [32] consists of breast ultrasound images, intended for binary classification. Originally based on a collection of 780 breast ultrasound images, the dataset is split into 546 training images, 78 validation images, and 156 test images. The original dataset categorizes images into three classes: normal, benign, and malignant. However, for simplicity, normal and benign cases are combined into a single positive class, and the task is framed as a binary classification problem where malignant cases are classified as negative. The original images of size 1 × 500 × 500 are resized to 1 × 28 × 28. The dataset evaluation metric is typically accuracy or AUC (Area Under the Curve).
3. NoduleMNIST3D: This lung nodule dataset collects images from thoracic lung nodule CT scans. The dataset is used in a benchmark paper [33]. Originally from [47], a total of 1,633 three-dimensional 28 × 28 × 28 images with spacing 1mm × 1mm × 1mm are cropped from the center of spatially normalized thoracic lung nodule scans. The data is turned into a binary classification task using malignancy levels 1 and 2 as negative, 4 and 5 as positive, ignoring malignancy level 3. With a splitting rate of 7:1:2, there are 1,158 training data, 165 validation data, and 310 testing data. The area under the curve (AUC) is used as an evaluation metric.

## 7.2 Training-free Metrics

The bottleneck of accelerating AutoML for medical problems is the costly and unstable DNN training, for each sampled DNN during the architecture search. To avoid such overhead, we aim to replace the DNN training with indicators or metrics that exhibit strong correlations with DNNs’ performance on healthcare benchmarks while not requiring training. Inspired by the recent development of deep learning theory [24, 27] and AutoML methods [20, 40], we introduce four training-free metrics. These four metrics are complementary to each other in characterizing different aspects of a DNN’s properties:

1. Condition number of Neural Tangent Kernel (NTK): Training deep networks requires optimizing high-dimensional non-convex loss functions. In practice, gradient descent often finds the global or good local minimum. However, many expressible networks are not easily learnable. For example, a deep stack of convolutional layers (e.g. Vgg [48]) is much harder to train than networks with skip connections (ResNet [38], DenseNet [49], etc.), even the former could equip a larger number of parameters. To characterize the training dynamics of wide networks, Neural tangent kernel (NTK) is proposed [50–52]. NTK controls the training dynamics of linearized DNNs and can be treated as the measurement of “sample-wise similarity”. NTK is defined as:

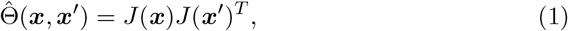

where *J* (***x***) is the Jacobian evaluated at input samples ***x***. Inspired by [27, 28], we measure the trainability of networks by studying the spectrum and conditioning of 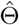, and use the empirical condition number of NTK to represent trainability:

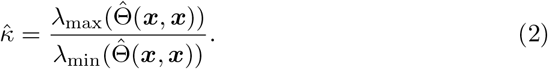 *x* are drawn from a training dataset. 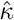 is calculated at the network’s initialization. A small NTK condition number indicates a smooth loss landscape and favors faster DNN training convergence.
  - Implementation: 1) We calculate each element of the NTK matrix as the inner product of network’s gradients across different input samples; 2) We calculate the eigenvalues of NTK, and then follow Equation 2.
  - Role: The trainability of a neural network studies how effective it can be optimized by gradient descent [53–55]. Since NTK controls the training dynamics of neural networks, its condition number can characterize the network’s trainability.
  - Reason to choose: Although trainability is only one property of deep networks, recent works still find that it is highly relevant to the final performance after networks are fully trained [28, 56, 57].
  - Length distortion (Length): In our work, we also study the manifold complexity of mapping the input through the network. We define the network as ℳ, its inputoutput Jacobian ***v***(*x*) = *∂*_*x*_ ℳ (*x*) at an input *x*. Length distortion measures the norm of a DNN’s input-output Jacobian. Since the ground-truth function we want to estimate (using ℳ) is usually very complex, one may expect that networks with better performance should generate longer outputs. We will calculate expected complexities over a certain number of *x*s randomly sampled. A large length distortion indicates a high sensitivity of DNN’s output to changes of its input, and thus potentially indicates a larger model capacity in learning complicated input-output mappings.
  - Implementation: We calculate ℒ^*E*^ via mapping inputs through the network. The Length Distortion in Euclidean space is defined as 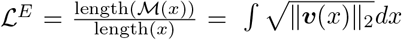 It measures when the network takes a unit-length input, what is the length of the output curve.
  - Role: We use ℒ^*E*^ to characterize the complexity of deep networks. Intuitively, a complex network can propagate a simple input into a complex manifold at its output layer, thus likely to possess a strong learning capacity.
  - Reason to choose: There are many ways to characterize network’s complexity, such as the number of model parameters, the number of linear regions [28], and norms of model weights [58]. However, the number and norm of weights are agnostic to model architectures. The complexity of computing the number of linear regions is high (quadratic to the number of input samples). In contrast, ℒ^*E*^ is very cheap to calculate (linear to the number of samples). Recent works show that ℒ^*E*^ strongly correlates to the network’s performance. [25]
2. Synaptic flow (SynFlow): This saliency criteria approximates the change in loss when a specific parameter is removed. Tanaka et al. [29] generalized these so-called synaptic saliency scores and proposed a modified version (SynFlow) which avoids layer collapse when performing parameter pruning. Synaptic flow measures the product of a DNN parameter and its gradient. Similar to NTK, we transform to the negative absolute value of SynFlow.

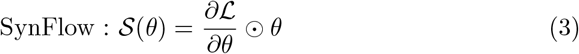 ℒ is the loss function of a neural network with parameters ***θ***, *θ ∈* ***θ***. *S* is the per-parameter saliency. We extend these saliency metrics to score an entire neural network by summing over all |***θ***| parameters in the model: 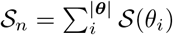
  - Implementation: We follow Equation 3, where we backward mini-batches of input through the network, and calculate the product between its parameters and their gradients.
  - Role: Synflow indicates the sensitivity of the loss with respect to changes in a specific DNN parameter (i.e. larger SynFlow indicates more important parameters). In general, at initialization, if the loss value is very sensitive to changes of model parameters, then it can be more easily optimized.
  - Reason to choose: Synflow [29, 59] is proposed to perform parameter pruning based on a saliency metric computed at initialization, and is found to be able to effectively indicate the importance of neurons.
3. Zero-shot Inverse Coefficient (ZiCo): In [31], Li et al. found that networks with high training convergence speed and generalization capacity should have high absolute mean values and low standard deviation values for the gradient with respect to the parameters across different training samples and batches. Therefore, ZiCo jointly considers both absolute mean and standard deviation values. ZiCo prefers high absolute mean values and low standard deviation values of a DNN’s sample-wise gradients. It implicitly improves the convergence and generalization of DNNs.

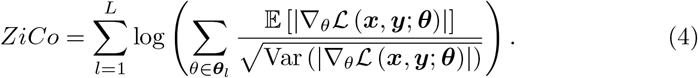 *L* stands for the depths of the network. ***θ*** denotes the set of all initialized parameters of a given network; ***θ***_*l*_ denote the parameters of the *l*^*th*^ layer of the network, and *θ* represents each element in ***θ***_*l*_; ***x*** and ***y*** are input samples and corresponding labels from the training set. 𝔼 and Var are taken over the batch of input samples.
  - Implementation: We follow Equation 4 to backward batches of input samples through the network, then calculate the mean and variance of gradients of each neuron.
  - Role: ZiCo is proposed to characterize both training convergence speed and generalization capacity.
  - Reason to choose: since ZiCo jointly considers two properties of deep networks (trainability and generalization), in [31] ZiCo was shown to achieve a strong selection of high-quality network architectures.

Instead of calculating the loss or error of a neural network after training, we measure these training-free metrics at the network’s initialization to characterize its performance. The core benefit of our training-free metrics is to achieve a strong correlation with the quality of designed networks with minimal computation costs. Note that for all four metrics, we use their expectations taken over batches of samples drawn from a training dataset ***x*** ~*D*_train_ and random Kaiming normal initializations 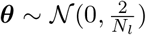 (*N*_*l*_ is the width at layer *l*) [60].

## 7.3 Model Space

We build our model space based on NAS-Bench-201 [34]. NAS-Bench-201 provides a standard cell-based search space (containing 15,625 architectures). The macro skeleton of each architecture candidate begins with a convolution layer as a stem, followed by three blocks of cells with two intermediate residual blocks connecting them. Then, global average pooling is applied to flatten the output into one dimension, followed by a fully connected layer for binary or multi-classification output, depending on the application. Each block contains 5 cells, with each cell having 4 nodes (feature maps) that form a directed acyclic graph, where each edge is associated with an operation selected from *none* (*zero*), *skip connection, conv* 1 × 1, *conv* 3 × 3, and *average pooling* 3 × 3, resulting in a total of 5^6^ = 15, 625 candidates. We adapt it to our problems by changing the convolution and pooling layer to 1D and 3D kernels. To obtain the benchmark results, we sample 150 architectures, which is approximately 150*/*15, 625 ≈ 1%.

## 7.4 Training-free Metrics Combination

To improve the robustness of training-free metrics across different clinical datasets, we seek to ensemble different combinations of the four training-free metrics. We employ a bootstrap method (*B* = 2000) over the 150 benchmark data points from the two distinct datasets. Within each bootstrap iteration, a 80 : 20 train-test split is applied separately on both the ECG2017 and NoduleMNIST3D datasets. Then, after merging training sets from both datasets, a multivariate linear ridge regression is conducted to compute the weighted aggregate of training-free metrics. The multivariate regression regresses the logarithm of all four training-free metrics to the standardized model test performance of both datasets.

To further explore the potential of using a reduced number of training-free metrics, we repeat the bootstrap procedure while retaining only three out of the four training-free metrics. Spearman correlation coefficients between the model performance and weighted aggregate of training-free metrics across ECG2017 and NoduleMNIST3D test sets are utilized to evaluate performance consistency. Because the rank correlation remains unchanged for linear rescale of the combined training-free metric, for simplicity, we divide the weights derived from the multivariate linear ridge regression by the minimum absolute coefficient to achieve a more interpretable overall proportion.

## 7.5 Architecture Search Methods

The overall pipeline of our method works as a modular framework of evaluation phase and search phase. Given a large model space (which includes over 10K different DNN structures in our work), we equip popular neural architecture search algorithms with our training-free metrics, i.e., these search algorithms will use the TEACUP metric as a reward to evaluate architectures when they explore the model space. For each architecture explored by the search algorithm, we randomly initialized the network and calculated the four training-free metrics on the network’s initialization with mini-batches of training data (batch size= 8 with 4 mini-batches), without any gradient descent steps. That means our search is still data-dependent and metrics are aware of different tasks. The final searched network architecture will be evaluated by training.

### 7.5.1 Reinforcement Learning (RL)

The policy agent maintains an internal state to represent the architecture search space, denoted as ***θ*** ^***A***^. This internal state can be converted to a categorical distribution of the architectures (***𝒜***) via softmax: ***𝒜*** = *σ*(***θ***^***A***^).

#### Stopping Criterion

We stop the RL search when the average TEACUP metric stops increasing for 30 iterations (total iterations *T* = 100 in our work). We trained the RL agent with Adam optimizer and a learning rate as *η* = 0.001.

#### Architecture Sampling

In each iteration, the agent samples 10 architectures *a*_*t*_ from ***𝒜***.

#### Update

We update the RL agent via policy gradients.

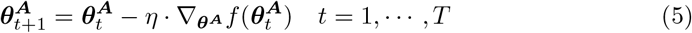

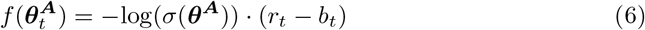

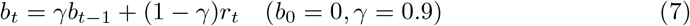

*r* stands for reward, the TEACUP metric combined in Sec. 7.4, and *b* for an exponential moving average of reward for variance reduction.

#### Architecture Deriving

To derive the final searched network, the agent chooses the architecture that has the highest probability, i.e., *a*^∗^ = argmax_*a*_*σ*(***θ***^***A***^)(*a*).

#### 7.5.2 Genetic Algorithm

We adopt a Bayesian probabilistic model building genetic algorithm as another search method. The genetic algorithm is first initialized with a population of 50 architectures by random sampling.

##### Stopping Criterion

We run the algorithm for a maximum of 100 iterations, and early-stop the searching when the average TEACUP metric stops increasing for 30 iterations.

##### Architecture Sampling

We model the distribution of the architectures ***𝒜*** as multinomial distributions

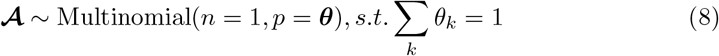

That is, we draw *n* = 1 instance each time from {1, 2, .., *k*, .., *K*} classes. The probability of each class *θ*_*k*_ follows a Dirichlet distribution as an uninformative prior:

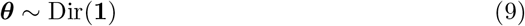

The posterior distribution of ***θ*** will be updated using the survived architectures in each generation that further updates the sampled architectures.

##### Update

Following Zhang et al. [35], in each iteration, the population is updated by adding a set of new architectures (n=10 in our work) and popping out the same amount of oldest architectures (the one that stays in the population for the longest time). Our buffer keeps at most 5 sets of architectures, which is 50 architectures in total. At the end of each iteration, we calculate the mean reward *r*_*mean*_ in the buffer and only use the *M* architectures with a higher reward than *r*_*mean*_ for updating the probability ***θ*** by generating the conjugate rule of Dirichlet-Multinomial:

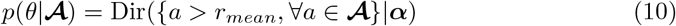

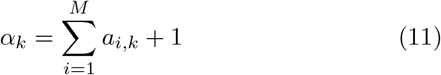

##### Architecture Deriving

To derive the final searched network, the best architecture from the population is selected, where the criterion is the same as we choose *a*_*t*_ (see above “Architecture Sampling”).

#### 7.5.3 Pruning

Inspired by recent works on pruning-from-scratch [59, 61], we also leverage a a pruning-by-importance model search mechanism [28] to quickly shrink the search possibilities and boost the search efficiency. Specifically, we start the search with a super-network *M*_0_ composed of all possible operators and edges, and iteratively prune operators with marginal importance (with respect to four training-free metrics). In the outer loop, for every round we prune one operator on each edge with the least contribution of four training-free metrics. In the inner loops, we measure and compare each operator’s importance.

##### Stopping Criterion and Architecture Deriving

The outer-loop stops when the current supernet *ℳ*_*t*_ is pruned to be a single-path network, i.e., there is only one candidate operator left on each edge.

##### Update

For the inner-loop, we measure the change of each four training-free metrics before and after pruning each individual operator, and assess its importance using the sum of ranks of the changes of four training-free metrics. We order all currently available operators in terms of their importance, and prune the lowest-importance operator on each edge.

For more details, please refer to the Algorithm 1 in [28].

### 7.6 Training Settings

We use a similar training setting as stated in the NAS-Bench-201[34] for both the benchmark 150 architectures and the evaluation of the optimized architecture generated from search methods. We use SGD and cosine annealing learning scheduler for all three datasets. When the early stop is adopted, we use the checkpoint that has the lowest validation loss as the final model. We summarize our training settings in Table 1.

### 7.7 Evaluation of Model Ensembles

We ensemble the predictions by voting and taking averages for ECG2017, BreastMNIST, and NoduleMNIST3D, respectively, based on their reward metrics (F1 for ECG2017, and AUC for BreastMNIST and NoduleMNIST3D). For ECG2017, since it is a multiclass classification with an F1 score as the reward, we set the majority vote among 4 classes as the final prediction. For BreastMNIST and NoduleMNIST3D, since they are binary classifications with AUC as the reward, we take the average over all the model outputs.

**Supplementary Table 1:** Training settings on three datasets.

| Dataset | LR | Momentum | Weight Decay | Epochs | Early Stop |
| --- | --- | --- | --- | --- | --- |
| ECG2017 | 0.01 | 0.9 | $1 \times 10^{-7}$ | 10 | Yes |
| BreastMNIST | 0.001 | 0.99 | 0.001 | 200 | No |
| NoduleMNIST3D | 0.1 | 0.9 | 0.005 | 100 | No |

To compare the disparities between ensemble over architecture-wise and initialization-wise predictions, we select the most common architecture from a pool of 30 optimized models obtained through all three searching algorithms. Subsequently, we train the architecture for 10 times employing different random model initializations to form a robust control group. We refer to this baseline as “single-architecture ensemble”, as opposed to ensembling over different architectures (referred to as “TEACUP ensemble”).

Then, we analyze the prediction uncertainties for each test case by ranking them based on the variance among the predictions generated from different architectures or initializations. The test case exhibiting the lowest variance across the ten predictions indicates the least uncertainty, and hence the highest prediction confidence.

Finally, we compute the accuracy for the test cases that ranked from up to the top 5% prediction confidence to the entire dataset (100%). The majority voting among the 10 predictions is used when computing accuracy to avoid significant tremors when calculating AUC on a relatively small amount of data points.

**Supplementary Table 2:** Training-free metric values, test rewards, and Spearman correlation coefficients across benchmark 150 architectures.

| Dataset | Content | Mean | Std | Length | NTK | SynFlow | ZiCo |
| --- | --- | --- | --- | --- | --- | --- | --- |
| **ECG2017** | Length | 93.28 | 296.41 |  |  |  |  |
| | NTK | $1.78 \times 10^9$ | $1.28 \times 10^{10}$ | 0.920 | | | |
|  | SynFlow | 1.53 | 0.31 | 0.591 | 0.567 |  |  |
|  | ZiCo | 304.97 | 146.18 | 0.477 | 0.452 | 0.170 |  |
|  | Test reward | 0.62 | 0.02 | 0.307 | 0.362 | 0.016 | 0.437 |
| **BreastMNIST** | Length | 784.00 | 2,450.00 |  |  |  |  |
| | NTK | $7.33 \times 10^7$ | $5.53 \times 10^8$ | 0.887 | | | |
|  | SynFlow | 8.87 | 1.37 | 0.247 | 0.219 |  |  |
|  | ZiCo | 330.00 | 158.00 | 0.470 | 0.421 | 0.050 |  |
|  | Test reward | 0.85 | 0.05 | 0.248 | 0.256 | 0.235 | 0.384 |
| **NoduleMNIST3D** | Length | 1179.89 | 3192.46 |  |  |  |  |
| | NTK | $1.07 \times 10^5$ | $7.36 \times 10^5$ | 0.866 | | | |
|  | SynFlow | 23.88 | 4.15 | 0.666 | 0.624 |  |  |
|  | ZiCo | 349.57 | 168.77 | 0.417 | 0.352 | 0.187 |  |
|  | Test reward | 0.85 | 0.03 | 0.435 | 0.487 | 0.265 | 0.063 |

**Supplementary Table 3:** Bootstrap results (*B* = 2000) across different combinations of training-free metrics. Using all four training-free metrics reaches an overall best result.

| Training-free metrics used | Spearman correlation with model performance |  |  |
| --- | --- | --- | --- |
|  | ECG2017 | NoduleMNIST3D | Summation |
| All (Length + NTK + SynFlow + ZiCo) | $0.41 \pm 0.14$ | $0.43 \pm 0.16$ | **0.84** |
| NTK + SynFlow + ZiCo | $0.38 \pm 0.15$ | $0.44 \pm 0.16$ | 0.82 |
| Length + SynFlow + ZiCo | $0.35 \pm 0.15$ | $0.28 \pm 0.18$ | 0.63 |
| Length + NTK + ZiCo | $0.45 \pm 0.13$ | $0.38 \pm 0.17$ | 0.83 |
| Length + NTK + SynFlow | $0.35 \pm 0.15$ | $0.48 \pm 0.15$ | 0.83 |

**Supplementary Figure 1:**
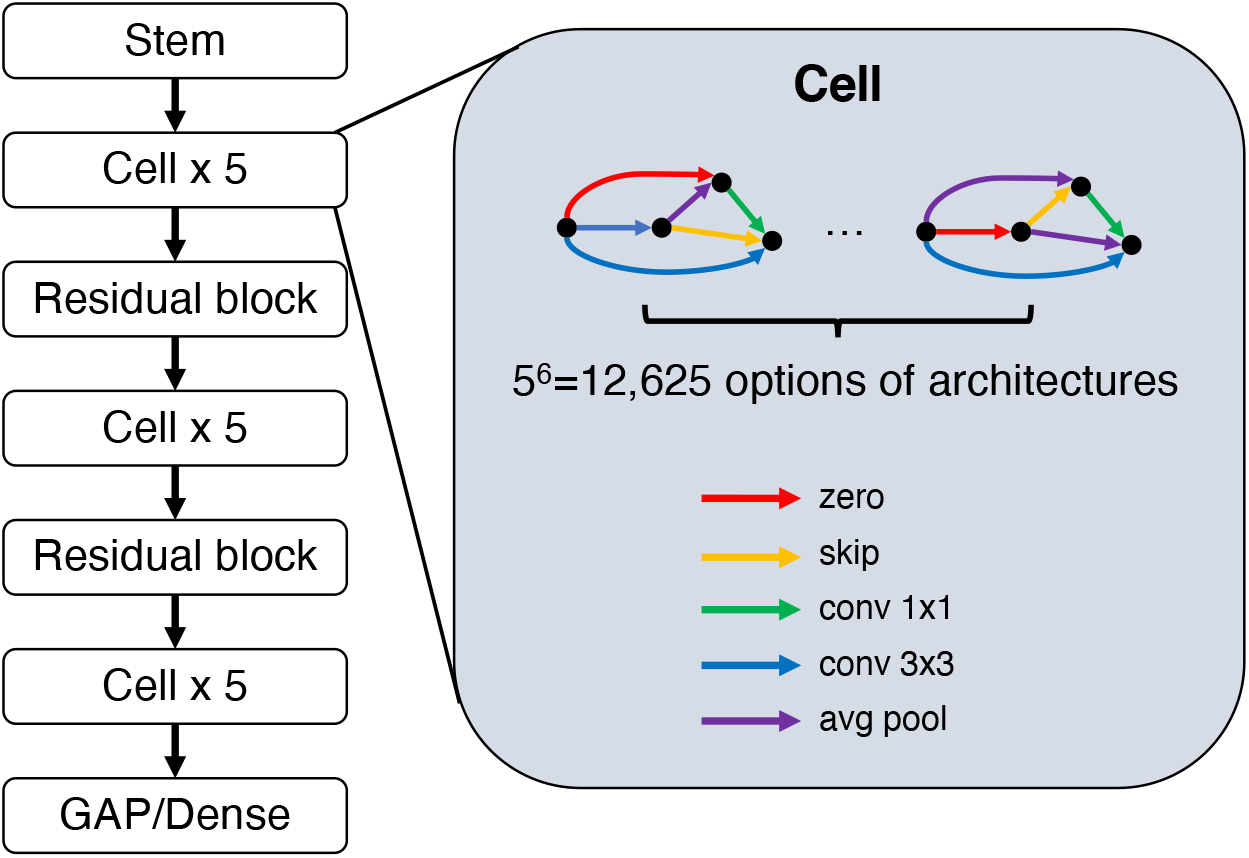
Illustration of NAS-Bench-201 model space.

**Supplementary Figure 2:**
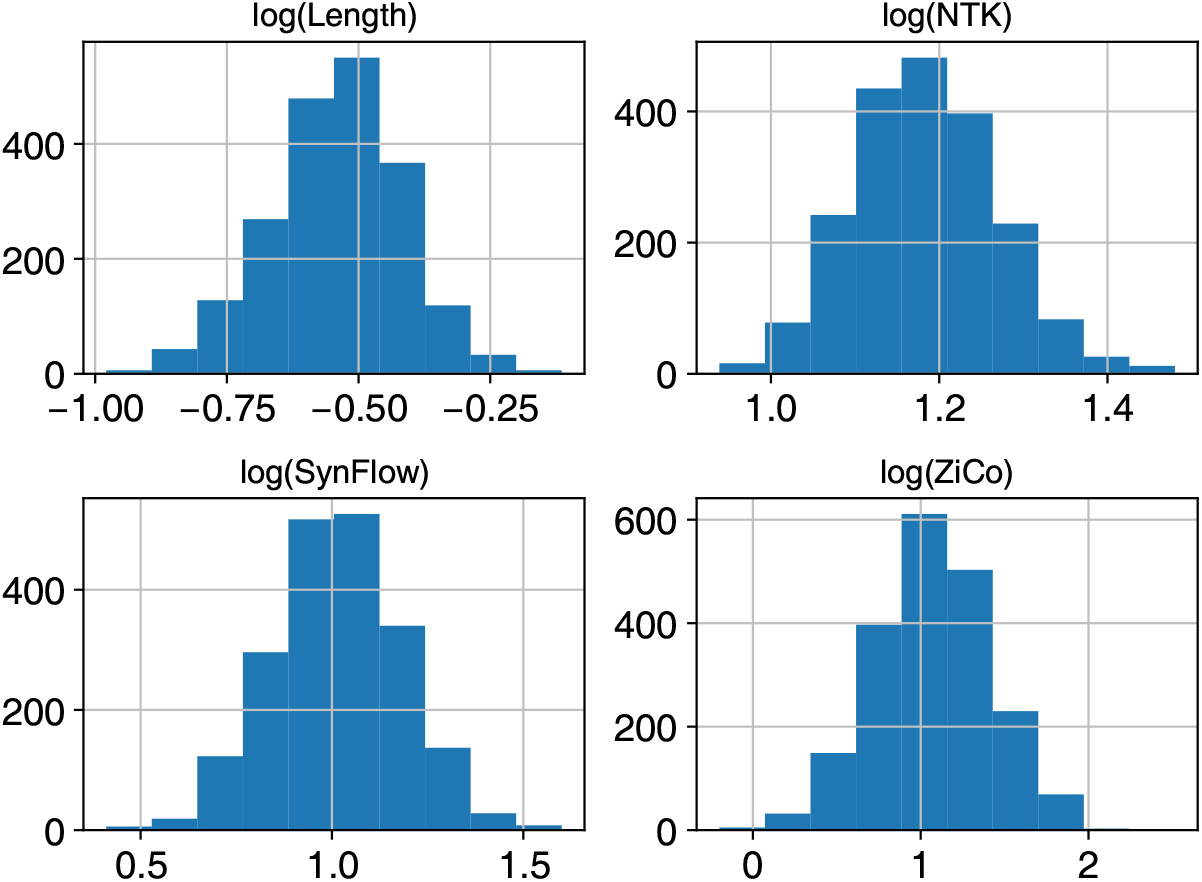
Bootstrap results of the coefficient distribution on the four log training metrics.

**Supplementary Figure 3:**
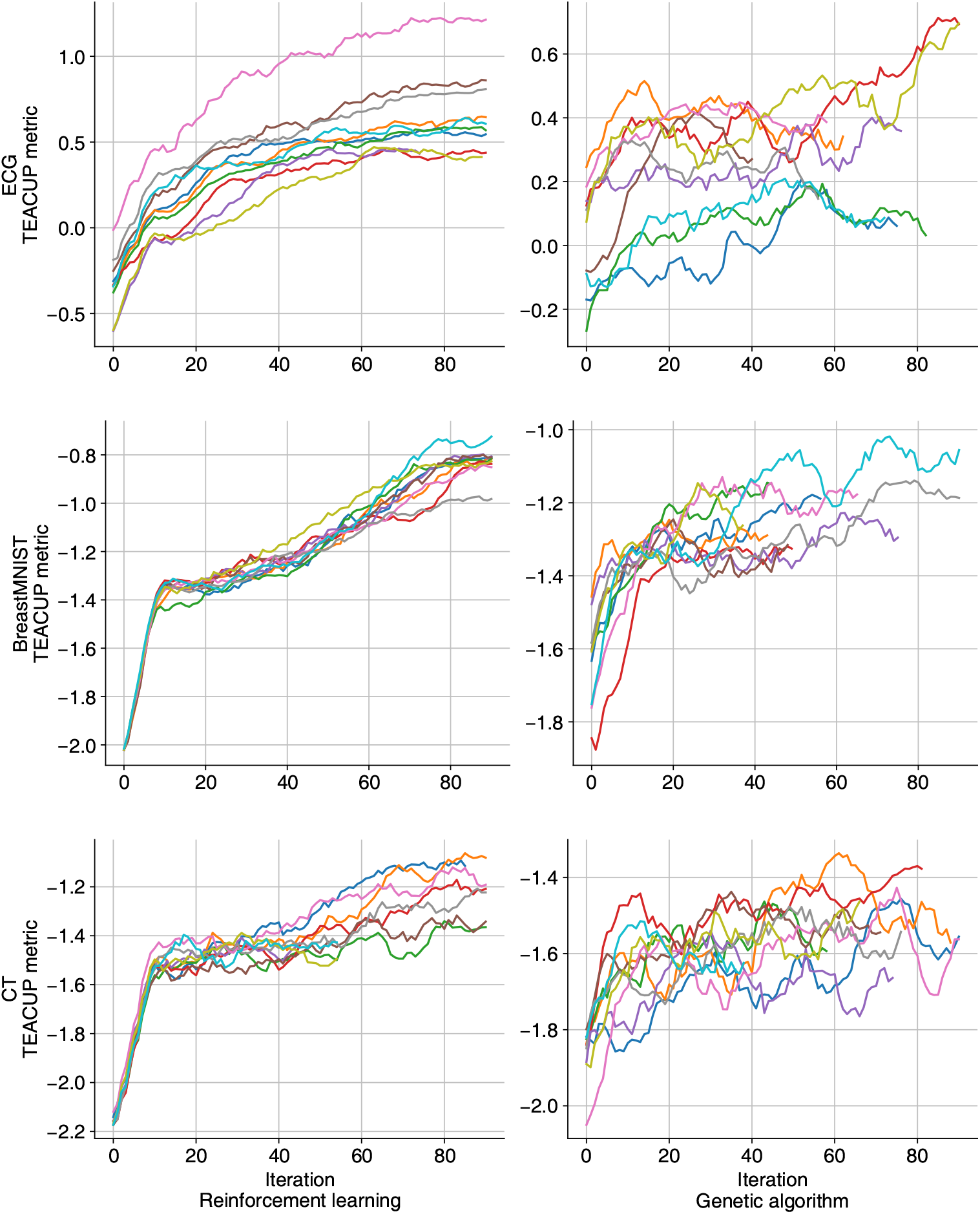
Change of TEACUP metrics as iteration increases in reinforcement learning and genetic algorithm. Different colors represent different, independent search runs

## Footnotes

1 We use one cloud A100 on AWS (https://aws.amazon.com/ec2/instance-types/p4/) as an example: $4.1 per hour *×* 0.14 hours per architecture *×* 12800 architectures ≈ $7347.

